# The Development of a Care Home Data Platform in Scotland: Insights from the Care Home Innovation Partnership, Lothian

**DOI:** 10.1101/2020.08.17.20176503

**Authors:** Lucy Johnston, Jo Hockley, David AG Henderson, Susan D Shenkin

**Author notes:** This work was funded by Health Innovation South East Scotland in collaboration with the Chief Scientist’s Office. We would like to acknowledge the contribution of our care home partners in CHIP.

## Abstract

Care homes collect a large amount of data about their residents, and the care provided, but there is a lack of consistency in how this information is collected. There is also a need to minimise the burden of data collection on staff, ensure information informs and supports person-centred care, and that this data is then of use to regulatory agencies, policy makers and researchers.

We examined the data collected in six Care Homes in Lothian, Scotland. We extracted the meta-data collected, cross-referenced definitions and assessed the degree of current harmonisation between individual care homes and with data sets currently in use in Scotland and internationally. We interviewed the care home managers to identify data collection processes, views and experiences of current data availability, gaps, access and issues of capacity and capability in relation to data management and analytics.

Our work has illustrated the scale of the data collected by care homes, the varied formats and heterogeneity of scope and definition. The inventory of 15 core data items that emerged, serves to expose in detail the *foundations* of care home data sets. The groundwork illuminated the heterogeneity in tools and assessments used to generate the data and the way in which the data is to be used, affects how it is specified and frequency of collection. By making known the reality of how and why care home data is collected, we can understand better the nuances of each individual data item that collectively create a data platform. We make four recommendations for the development of a national care home data platform.

## Introduction

In Scotland, over 35,000 older people live in around 850 care homes. Of these 58.6% are privately owned, 27.1% owned by voluntary or not-for-profit providers and 14.3% by local authority or Health Board. By 2040, it is predicted that 40% of people will live and subsequently die in a care home (Bone, et al. 2017). The need to make this a safe and sustainable place to live and be cared for is one of the most urgent challenges over the next 20 years. The scale and immediacy of this challenge has been underlined by the severe and evolving impact of the COVID-19 pandemic on residents (Burton, J. et al MEDRx, July 2020), their loved ones and the staff (McGilton, K. et al., 2020) who care for them. Rising to the challenge will require high-level strategic and operational responses informed by timely and accurate information.

Care homes collect a large amount of data to support the care of residents, and to report to regulatory authorities. Improved digital connectivity and flow of data within and between care homes and health and social care providers will enhance the quality of care for each resident, better evidence practice and quality improvements, and more robustly inform regional and national policies. Progress in these fields is all contingent on the curation of a robust data set, with agreed definitions and effective curation and access systems. However, at present, there is no standardisation of what is collected and data linkage is minimal (Hanratty, B. et al, 2020), International and UK programmes are in progress, which aim to develop common data elements (Corazzini, KN. et al., 2019) and a Minimum Data Set by the DACHA study (NIHR, 2019) for care homes.

The Covid-19 pandemic has laid bare the consequences of these data deficits and intensified activities to address these limitations (Hanratty, B. et al, 2020); the long-needed priority and attention now afforded to the subject is an opportunity to build better. There is no doubt this is a major challenge for the current system and structure which struggles even to identify accurately who lives in a care home (Burton, J. et al., 2019).

The groundwork considered necessary to inform the development of a data platform for care homes was conducted between July 2019 and January 2020. It aimed to establish what data are routinely collected as part of resident care in six different care homes and the degree of current harmonisation between the homes and with Scottish and international data sets. To achieve the aim, the following objectives were set: (1) Identify current care home data sets populated by care homes; (2) Create an inventory of data routinely used by six care homes and compare similarities with current data sets and (3) Collate care home views and experiences of current data availability, gaps and access issues.

## Methods

The six Lothian care homes involved in the study are members of the Lothian Care Home Innovation Partnership (CHIP Lothian). CHIP Lothian was formed in autumn 2018 as a result of workshops held across Lothian to update care home managers regarding the wider vision for a teaching/research-based care home centre for Lothian and South East Scotland (Hockley, J. et al., 2016).

The homes represent, independent, third sector and local authority managed care homes for older people (see Table 1). They also recorded information about their residents in a variety of forms: paper records, bespoke electronic data capture and/or commercial data platforms. Electronic Care Planning applications are used in five of the six partner care homes and one uses medication management software that links to their chosen care planning system. One home records all data within hard copy patient records and selected indicators are then entered into internal analytical packages (mostly excel).

**Table 1:** Care planning /data collection system in partner care homes

| Partner Home | No. Beds | Sector | Data Collection Packages |
| --- | --- | --- | --- |
| 1 | 72 | Independent | Caresys |
| 2 | 72 | Charity | iCare Health and e-med |
| 3 | 63 | Independent | Personcentred Software (PCS) |
| 4 | 70 | Charity | Personcentred Software (PCS) |
| 5 | 61 | Local Authority | Kardex and Internal Computer System |
| 6 | 80 | Independent | Paper based |

A Research Fellow (LJ) contacted the managers and arranged to visit the home to review and document meta data on individuals residents collected by the staff. The definitions and scope of each resulting data inventory were then compared and cross-referenced, to set out the degree of current harmonisation. Interviews with six ‘partner’ care home managers were also undertaken to identify data collection processes, views and experiences of current data availability, deficits and issues of capacity and capability in relation to data management and analytics. A further group discussion with these managers to ‘sense’ check the findings was held. A scan of the literature, focussing on known topic experts and discussions with key Scottish care home data stakeholders (users and analysts) were also undertaken. These were used to identify key current care home ‘data sets’ in use in Scotland and internationally. The Care Home data inventories were compared to these ‘data sets’ again to see the degree of harmonisation.

This work was conducted between July 2019 and January 2020. Ethical approval was given by the Edinburgh Napier University SHSC Ethics Committee (Ref/SHSC2007).

## Results

### Care Home Data Sets

Our groundwork identified five sources of care home data or data sets of relevance to the development of a national care home data platform in Scotland. Two established Scottish care home data sets were populated via the Social Care Survey (SCS) and Scottish Care Home Census. These have now been superseded by the SOURCE data collection curated by Public Health Scotland. In addition, a new COVID-19 specific data set on care homes is being generated via the ‘Safety Huddle Template’. Care home data sets have also been constructed (predominantly in European countries, America and New Zealand) through the use of The Resident Assessment Instrument-Minimum Data (RAI-MDS) and the International Prevalence Measurement of Care Problems in Care Homes (Landelijke Prevalentiemeting Zorgkwaliteit, or LPZ for short, from the Dutch).

### The Social Care Survey (SCS) and Scottish Care Home Census

Historically, there were two national data sources relevant to care home residents – The Social Care Survey (SCS) and Scottish Care Home Census. The exact content of these sources and their potential strengths and limitations are detailed elsewhere (Henderson, DAG, et al., 2019). However, as Table 2 shows, data harmonisation has not been achieved for basic profile data on residents. Care Homes made an annual ‘data return’ to these sources and a number of features of the SCS and SCHC may undermine the foundations of a national data platform. For example, data returns were not mandatory for SCHC and therefore not complete and a quality assurance process was lacking. In addition, neither source collected any clinical measures or key diagnosis but recorded only broad clinical variables (SCHC) and client group category (SCS).

**Table 2:**
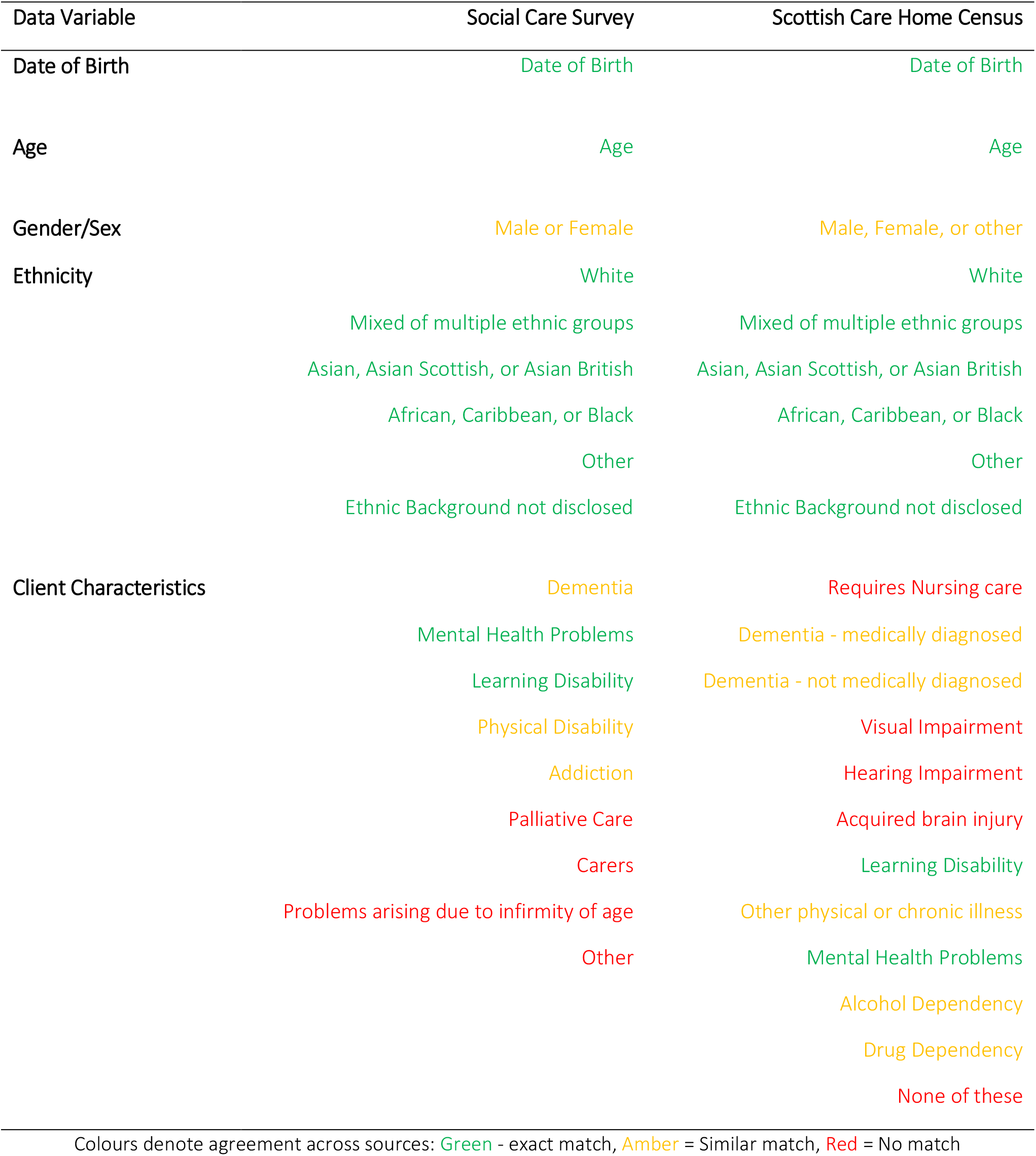
Comparison of SCS and SCHC demographic categories

Whilst SOURCE addresses some of the issues described above, it contains less information about care homes. Furthermore, the timeliness of publication is tardy. To date only an experimental statistical release for financial year 2017/18 has been published (Information Services Division, 2019). As of August 2020, data collected for financial year 2018/19 have, not yet been analysed in sufficient form for public release.

### NHS Education Scotland /NES Digital – Safety Huddle Template

On 17 May 2020, the Scottish Government Cabinet Secretary for Health and Sport announced Coronavirus (COVID 19): enhanced professional clinical and care oversight of care homes (Scottish Government, 2020a) as a result of the Coronavirus pandemic (COVID 19). This set out the expectation that care homes would populate a Safety Huddle Template to provide COVID-19 specific and related workforce information including for example outcomes of testing. Additional information to aid staffing decision-making was specified as:

- Number requiring 1:1 care
- Number receiving End of Life Care
- Number of residents with significant cognitive impairment
- Number of residents transferred to hospital (non-Covid related)

A digital version of the Template was made available in August 2020 (Scottish Government 2020b) to assist care homes complete the template, however it is not mandatory to do so. Under consideration for future versions of the Safety Huddle Template is the inclusion of data for indicators based largely on Care Inspectorate returns/e-form). The proposed data on residents as at 3 June 2020 are listed below.

- Fluid Food Nutrition/ Malnutrition Universal Screening Tool (MUST Score)
- 2 or more co-morbidities
- Frailty score greater than or equal to 7
- falls with harm
- new pressure ulcers Grade 2 and above
- medication errors
- residents with Advance Care Plan
- residents with Do not attempt cardio-pulmonary resuscitation (DNACPR)

### The Resident Assessment Instrument-Minimum Data (RAI-MDS)

The RAI-MDS is a standardised assessment tool to “collect the minimum amount of data to guide care planning and monitoring for residents” (Hutchinson, A., et al. 2010). It includes a core set of data on specified prevalence and incidence across a number of domains for each assessed individual. Its use and development is guided by an international consortium of researchers and clinicians from over 30 countries, known as the interRAI network (http://www.interrai.org). This data set is generated mostly by care homes in mainly northern Europe and America, Asia and the Pacific Rim. In New Zealand use of the interRAI assessments are mandatory nationwide in all care settings and are conducted in a single software platform.

### LPZ – The International Prevalence Measurement of Care Problems in Care Homes

LPZ is operated as an annual measurement of the quality of care provided by subscribing institutions. Six Care Indicators are included in the measurement, with institutions providing data on one or any number of the indicators, depending on the local focus of quality improvement. Five of the LPZ Care Indicators overlap to a large degree with those specified by RAI-MDS. In 2016 LPZ added the Care Indicator of ‘pain’ to its suite of measurements. In the UK only around 65 care homes currently use LPZ, mainly in the East Midlands (LPZ Network website).

The data specified by the RAI-MDS and LPZ is set out in Table 3.

**Table 3:** Data specified by RAI-MDS 2.0 Quality Indicators and LPZ Care Indicators

| Domain | RAI-MDS Indicator | LPZ Care Indicator |
| --- | --- | --- |
| Accidents | 1. Incidence of new fractures<br>2. Prevalence of falls | Falls |
| Behavioural and emotional patterns | 1. Prevalence of behavioural symptoms affecting others<br>2. Prevalence of symptoms of depression<br>3. Prevalence of symptoms of depression without ant-depressant therapy |  |
| Clinical Management | Use of nine or more medications |  |
| Cognitive Patterns | Incidence of cognitive impairment |  |
| Elimination and continence | 1. Prevalence of bladder/bowel incontinence<br>2. Prevalence of indwelling catheters<br>3. Prevalence of faecal impaction | Incontinence (urine, faecal, double) |
| Infection Control | Prevalence of urinary tract infections |  |
| Nutrition and eating | 1. Prevalence of weight loss<br>2. Prevalence of tube feeding<br>3. Prevalence of dehydration | Malnutrition |
| Physical functioning | 1. Prevalence of bedfast residents<br>2. Incidence of decline in late-loss ADLs<br>3. Incidence of declined in range-of-motion |  |
| Psychotropic drug use | 1. Prevalence of antipsychotic use in the absence of psychotic and related conditions<br>2. Prevalence of anxiety/hypnotic use<br>3. Prevalence of hypnotic use more than two times in the last week |  |
| Quality of life | 1. Prevalence of daily physical restraints<br>2. Prevalence of little or no activity | Physical restraints |
| Skin care | Prevalence of stage 1-4 pressure ulcers | Pressure ulcers |
| Pain |  | Pain |

### Care Home Data Inventory

#### Data Domains, Source and Uses

Our scoping work allowed us to structure the data inventory collected about individual residents within three key domains: Demographics, Diagnoses and Detailed Care Planning Information (DCPI). These three data domains are the foundations of any developing data platform.

Within all care homes key demographic data on residents is collected on admission (for example age, gender etc) and high-level information on medical diagnosis and long term conditions (for example cancer, stroke, *dementia, diabetes etc*). We saw in Table 1, that it is much of this high-level data that populates national data sources annually.

However, DCPI is gathered in significantly more detail via the ongoing assessment and review of individual’s, functioning and care needs (physical and emotional). DCPI data is gathered first on admission and used to populate an individual’s care plan. Individual components of this overall ‘plan’ and therefore individual data points are reviewed, edited, updated in line with the homes policies, care practices and the cycle of regulatory reporting demands. As a result, some ‘data items’ can be recorded daily, others weekly, monthly or quarterly – and others once a year for regulatory compliance returns. Data will be updated or amended when specific aspects of functioning and care provided are re-assessed. For example in response to a change in the condition of the resident.

Assessment, and as a result DCPI, is therefore multi-faceted, wide-ranging and fluid. In most care homes 15 plus separate aspects relating to each individual can and are assessed. To illustrate the scale and the variability between care homes, Box 1 lists the separate assessments used by two Care Homes to construct an individual resident’s care plan. They are listed in alphabetical order and not all assessments are included.

The scoping found that whilst the focus of individual assessments are similar, as with the national data sources and proposed indicators, they are not standardised and therefore not comparable.

##### Box 1: Examples of the number and range of assessment components for care planning in two care homes

| Example A | Example B |
| --- | --- |
| Acute Infection Care Plan | Behaviour |
| Anticipatory Care Planning/End of Life Care | Bladder & Bowel |
| Breathing and Smoking | Choking |
| Bowel & Bladder | Communication |
| Communication | Dependency Rating Scale |
| Eating & Drinking | DNACPR/End of life decisions |
| Falls | Eating & Drinking |
| Hearing/Vision | Falls Risk |
| Life History/This is me | Mood |
| Medication and Treatment | Oral Health |
| Mental Health, Cognition,<br>(Memory/Understanding), and Personal Safety | Personal Care |
| Mobility | Waterlow Risk |
| Oral & Dental |  |
| Personal Hygiene |  |
| Restraint Risk |  |
| Sleep & Resting |  |
| Waterlow/Pressure Ulcer Assessment |  |

Within a care home, the frequency of assessments and the extent to which key indicators are collated are driven by the needs and demands of those who want to use the data. Who uses the data and for what purpose determines for example what is routinely collected as opposed to what is regularly collected by individual homes. The way in which the data use and user impacts on the source has to be understood. Box 2 summarises the main reasons care managers gave for collecting the data items they do.

##### Box 2: Rationales for data collection by individual care homes

- Alerting care staff to a need for new/different care i.e. a change in resident’s condition/preferences
- Informing/sharing intelligence internally and externally – among staff (shift handovers for example) and with external care partners (for example care coordination and care transitions)
- Recording and evidencing care in the home, resident care pathways, transitions and use of external NHS resources
- Evaluating care – providing source evidence for outcomes and indicators of care
- Complying with regulations/governance/inspection
- Managing workforce and budget
- The COVID-19 Pandemic has added a further rationale for data

## Inventory

The data or information <u>about residents</u> most <u>***regularly or routinely***</u> collected and recorded by care homes is listed below.

1. Dependency/indicator of need score
2. Nutrition
3. Weight of resident
4. Incidence of falls
5. Incidence of pressure sores
6. Prevalence of infections
7. Wounds (new and ongoing)
8. Frailty
9. Bowel Movement(s)
10. Fluid intake
11. Mood/behaviour (delirium/depression)
12. Pain
13. Movement
14. Sleep
15. Observations/ Vital Signs (e.g. Systolic Blood Pressure (mmHg); Diastolic Blood Pressure (mmHg); Heart rate; Oxygen Saturation (%); Temperature; Blood Glucose Level (mmol/L); Respiration (Breaths per minute?)

Table 4 below represents this inventory of 15 data items alongside the assessment tools and measures used by each of the six care homes.

**Table 4:** Data sources – Assessment tools/measures for each data inventory item

| Inventory No. | Area assessed | 1 | 2 | 3 | 4 | 5 | 6 |
| --- | --- | --- | --- | --- | --- | --- | --- |
| 1 | Dependency/<br>indicator of<br>need | Augmented<br>IoRN | IoRN | Dependency<br>assessment | IoRN | IoRN | Organisation<br>Form |
| 2 | Nutrition | MUST | MUST | MUST | MUST | Eating Well<br>in Care homes/<br>Cook Safe | Organisation<br>Form |
| 3 | Weight | Kg/BMI | Kg/BMI | Kg/BMI | Kg/BMI | Kg/BMI | Kg/BMI |
| 4 | Incidence and<br>risk of falls | FRASE | Organisation<br>Form | Falls Risk | Falls Risk | Organisation<br>Form | Organisation<br>Form |
| 5 | Incidence and<br>risk of pressure<br>sores | Braden | Pressure Ulcer<br>Cross/<br>PU Checklist/<br>Waterlow | Waterlow | Waterlow | Waterlow | Skin integrity<br>Care Plan |
| 6 | Infections | Count/<br>type of infection | Count/<br>type of infection | Count/<br>type of infection | Count/<br>type of infection | Count/<br>type of infection | Count/<br>type of infection |
| 7 | Wounds<br>(new and<br>ongoing) | Internal<br>Chart | STAR<br>Classification | Wounds<br>assessment | Chart<br>on PCS | NHS Wound<br>Assessment Chart | Organisation<br>Form |
| 8 | Frailty | CIRC | SPAR Tool | Edmonton Frailty<br>Scale | Clinical Frailty<br>Scale | Not collected | Not collected |
| 9 | Bowel<br>Movement(s) | Bristol Stool<br>Chart | Bristol Stool<br>Chart | Chart<br>on PCS | Bristol Stool<br>Chart | Bristol Stool<br>Chart | Organisation<br>Form |
| 10 | Fluid<br>intake | Internal<br>Chart | Organisation<br>Form | Chart<br>on PCS | Chart<br>on PCS | Organisation<br>Form | Organisation<br>Form |
| 11(a) | Mood:<br>Depression | Geriatric<br>Depression scale/<br>Cornell scale for<br>Depression in<br>dementia | No measure/tool<br>reported* | No measure/tool<br>reported | Cornell scale for<br>depression in<br>dementia | No measure/tool<br>reported | No measure/tool<br>reported |
| 11(b) | Mood:<br>Delirium | 4AT | 4AT | Not recorded | Not recorded | Care support plan | Not collected |
| 12 | Pain | PAINAD<br>Doloplus2<br>Abbey Pain Scale | Abbey Pain Scale | Abbey Pain Scale | Abbey Pain Scale | Abbey Pain Scale | Abbey Pain Scale |
| 13 | Movement | Roper, Logan,<br>Tierney model of<br>ADL<br>Nolan's 6 senses<br>f/work | Care Support<br>Plan | No measure/tool<br>reported | No measure/tool<br>reported | Care Support<br>Plan | In Care<br>Plan |
| 14 | Sleep |  | Care Support<br>Plan | Care Support<br>Plan | Organisation<br>Form | Organisation<br>Form | Organisation<br>Form |
| 15 | Observations/<br>Vital Signs | ←----- Various charts -----→ |  |  |  |  |  |

The diversity and range of assessment tools and measures is a key limitation for data users. Compounding the differing ways in which data is specified and collected, across the homes the regularity or frequency of the recording of data on these items is also heterogeneous. Some for example record data on bowel movements of a resident only when their condition/health indicates this was needed to help assess and determine any additional care needs. In other homes, such data is recorded about every resident daily.

All care homes record and report falls, however specification of even this widely used data item is not universally standardised. Falls could, for example, be viewed and therefore recorded differently depending on the way in which the data was to be used. That is differently, if reporting for regulatory reasons, from manager intelligence who may want to differentiate between for example ‘a witnessed fall’ from an ‘unseen fall or indeed the severity of the fall – where ‘a fall with injury’ or ‘a fall resulting in hospitalisation’ would be recorded separately for one user, but combined for another.

In our inventory the data items numbered 10 to 15 are viewed by care homes as intelligence vital to monitoring a residents changing or deteriorating condition. That said, with the exception of number 15 – Observations, the other four to a large part rely on observation, care staff intuition and an assortment of ways to measure.

Further reinforcing the need to appreciate the users and source of data items in care homes is the need for detail within the high level meta data. Six very specific data items from within this inventory are viewed by managers as key signs of resident’s health, the quality of their care and the need for additional or enhanced care. They saw the number of residents with urinary or chest infections or losing weight as important data. In addition, the prevalence of pressure sores, falls and incidents of ‘challenging behaviour’ are closely monitored and therefore recorded as some form of ‘data’ in most homes daily.

The need to first stabilise the foundations before proceeding to develop a care home data platform is reinforced further by the results of cross-referencing our inventory with the data items being used by others in significant numbers. Table 5 illustrates the degree to which the inventory can be mapped across to the data elements used by MDS-RAI and LPZ. As can be seen, there are high level similarities but this is deceptive – at heart they are merely variations on a theme. The level of variation between care homes and across proposed data sets, unless harmonised and standardised can only serve to destabilise the much-needed data platform.

**Table 5:**
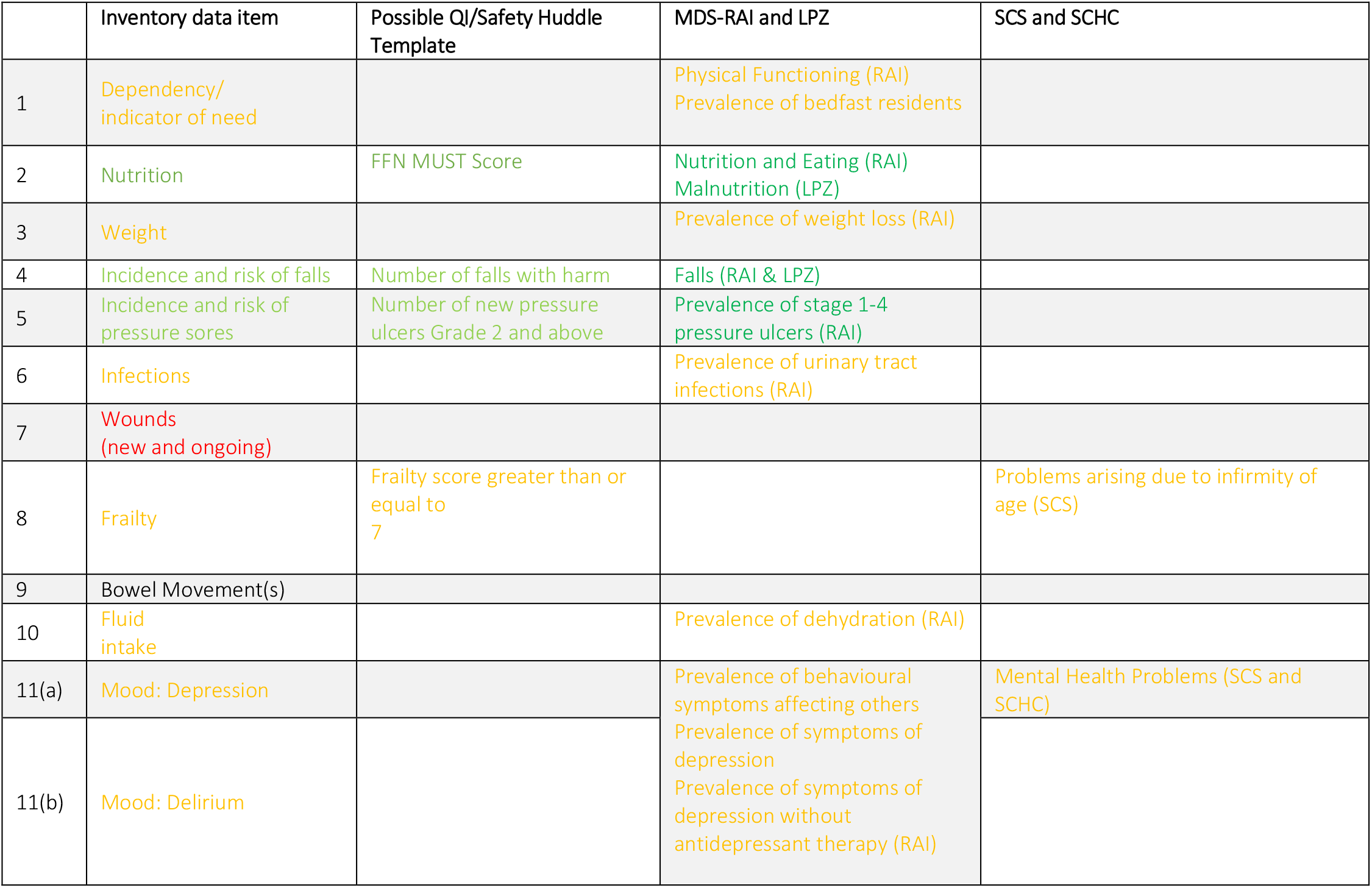
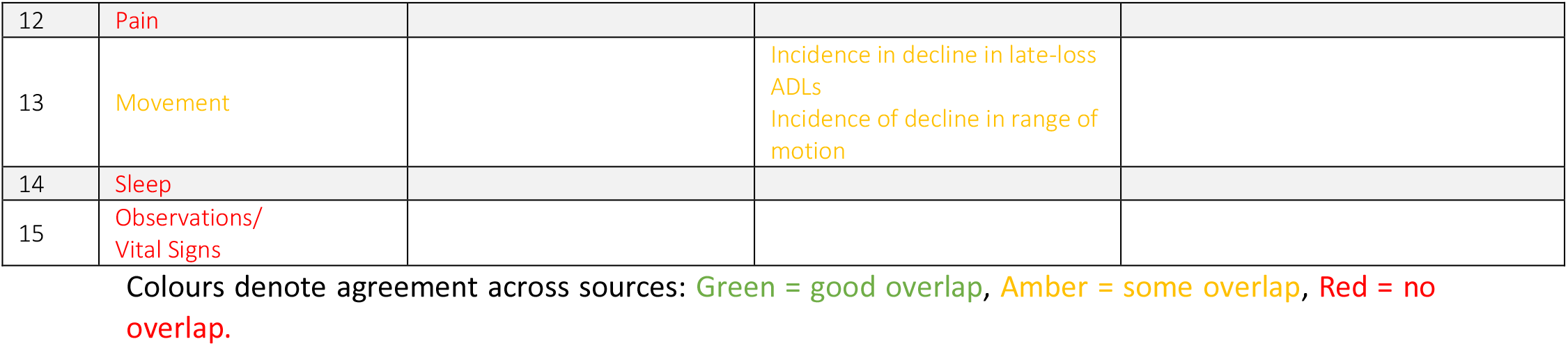
Extent of harmonisation across inventory and data sets

## Implications and Conclusions

Our work has illustrated the scale of the data collected by care homes, the varied formats and heterogeneity of scope and definition. An inventory of 15 core data items emerged from our work and exposed in detail the *foundations* of care home data sets. Whilst presented as a list, this straightforward numbering of data items belies the complexity and context specific nature of care home data. Refining and redefining data items will not be sufficient, unless the reality of data collection in care homes is addressed.

Our groundwork illuminated the heterogeneity in tools and assessments used to generate the data. It also highlighted that the way in which the data is to be used can affect how it is specified and frequency of collection. Most importantly, we have made explicit the reality of how care home data is collected, and the resulting need to understand the nuances of each individual data item. The content of national care home data sets are constructed via the specification, collection and reporting processes of local individual care homes. The inherent limitations, deficiencies and consequences of this are insufficiently understood. We recommend that work to improve specification, collection, curation and analysis in this area is best served by firstly investing in work to stabilise the foundations of care home data.

To comprehensively enhance the provision, flow and analysis of care home data items – the content of data sets must build *up* from what care homes tell us is of most importance to their work and therefore regularly gathered and used. Construction of a data platform must therefore also understand the capacity and capabilities of individual homes to provide useable data, and encompass the ‘needs’ of data users – inside and external to the home itself. Enhancements and improvements to data collection must reflect the culture of care and overall context of the home environment and not drive it. The priority being given to the recording of clinically related data and on care work completed being monitored can create a task orientated culture. It can also result in an over-reliance on more readily quantified aspects of care and every resident’s health and wellbeing. We recommend wider consultation with key stakeholders to determine ‘data needs’. There may be potential in Scotland proactively linking with the ongoing DACHA project to do this.

Two additional factors require attention to ensure they do not continue to undermine efforts to enhance and harmonise.

1. Data Linking and interoperability are extremely limited. As a result, the data can only function as stand-alone pieces of intelligence – both within and outside the home. The potential for better data analytics to support care decisions and inform wider policy from scrutiny or offer Incidence in decline in late-loss ADLs Incidence of decline in range of motion Sleep patterns and insights is restricted. Furthermore, the flow of information from care homes to care partners is ad hoc, lacks real-time capabilities and there are few examples of consistent interoperability. We recommend that a whole systems approach is taken to the development of a data platform for care homes in Scotland.
2. There is a need to question, that in the pursuit of the ultimate core data set or MDS – we ensure we do not compromise or limit what is needed to tell the full story of care homes. Too narrow a pursuit of the measurable or quantifiable, will lesson focus on meaningful process and outcome data that is by its nature soft data/qualitative data. For example, pre-scripted data fields limit the recording of social/emotional activities and related care provision, and the lived-experience of residents and causal factors relevant to resident outcomes are under recorded and less well understood. We recommend that a truly robust data platform for care homes must be co-produced – not by policy makers and sector representatives – but collectively, ensuring important insights form residents and frontline care staff inform the work.

## Data Availability

Source data is available. Please contact the authors

